# Night shift work and breast cancer incidence among women: a population-based cohort study

**DOI:** 10.1101/2025.09.02.25334911

**Authors:** Joy Sudan, Drinbardha Elshani, Arnaud Chiolero, Stéphane Cullati, Bernadette W.A. van der Linden

## Abstract

**Background:** Night shift work may increase breast cancer incidence, but evidence is inconclusive. We investigated the effect of night shift work on breast cancer incidence, considering life course night shift work characteristics, sleep and menopausal status.

**Methods:** We analyzed two UK Biobank cohorts, using Cox proportional hazards regression models. The first cohort (20,860 participants) investigated the effect of night shift work on breast cancer incidence (688 cases) from baseline (2006–2010) to the end of follow-up (2022), stratified by sleep patterns and menopausal status. The second cohort (64,539 participants) investigated night shift work characteristics (duration, frequency, length, consecutive shifts, rest days) and breast cancer incidence (4,455 cases), using retrospective data from participants’ first job to the end of follow-up.

**Results:** No effect of night shift work on breast cancer incidence was observed in the first cohort (hazard ratio (HR) 0.95, 95% CI: 0.85-1.06) or in the second cohort (HR 1.05% CI: 0.96-1.13). Stratification by sleep patterns and menopausal status showed no modifying effect. No night shift work characteristics were linked to breast cancer incidence.

**Conclusions:** Our findings suggest no effect of night shift work on breast cancer incidence, even when considering sleep patterns, menopausal status, and detailed life course characteristics.

## Introduction

Breast cancer is the most common cancer among women, with a global lifetime risk of 12% (1). Established risk factors include reproductive factors such as nulliparity and late menopause (2), lifestyle factors including alcohol consumption (3) and sedentary behavior (4), along with genetic mutations (5). Recent research has also identified disturbances in circadian rhythms, particularly those caused by night shift work, as an emerging risk factor for breast cancer (6,7). In many countries, night shift work has become increasingly prevalent due to the demands of a 24/7 economy, with 14% of women working night shifts in the European Union in 2020 (8).

Estrogen dysregulation due to melatonin desynchronization is a proposed mechanism linking night shift work to breast cancer. Melatonin not only regulates the sleep-wake cycle but also has well-documented anti-estrogenic and anti-tumor effects (9). When melatonin production is disrupted due to irregular sleep patterns (10), it may result in an imbalance of estrogen levels, a known risk factor for breast cancer (11). Factors such as sleep duration, sleep quality, and chronotype can influence how the body adapts to night shifts, thereby modulating the risk of estrogen-related breast cancer (12,13). Finally, the impact of night shift work on hormonal regulation may differ between premenopausal and postmenopausal women, as estrogen levels naturally vary between these groups, potentially influencing breast cancer incidence (6,14,15).

In 2019, the International Agency for Research on Cancer (IARC) classified night shift work, defined as work during the typical sleeping hours of the general population, as a probable human carcinogen. This classification was based on evidence from experimental animal studies and mechanistic insights, although direct human evidence remains limited (16). Indeed, the current evidence on the association between night shift work and breast cancer shows inconsistent results, even across meta-analyses (14,17–23). Case-control studies tend to report a positive association between night shift work and breast cancer incidence (12,15,24,25), while cohort studies do not support this link (26–28). The 2011 Canadian Burden of Occupational Cancer study estimated that between 2% and 5% of breast cancer cases in Canada can be attributable to night shift work (29).

A recurring limitation in the literature is the binary classification of night shift work, which typically compares breast cancer incidence between night shift workers and non-night shift workers at a single timepoint. This approach fails to consider the dynamic nature of night shift work characteristics throughout the working life course, such as monthly exposure frequency, number of consecutive night shifts, and rest periods (30). Few studies have examined those characteristics in detail, but those that have done so reported conflicting results regarding the effect of duration (18,19,31), frequency (14,15) and number of consecutive nights worked (32,33). Further research is required that takes into account the specific characteristics of night shift work over the course of an individual’s entire career.

Our goal was to assess the effect of night shift work on breast cancer incidence in women, using data from a large population-based cohort study. Specifically, we examined (1) the effect of night shift work on breast cancer incidence, and whether this effect was moderated by (2) sleep patterns (duration, quality, and chronotype) and (3) menopausal status. Additionally, we investigated (4) whether night shift work characteristics across the life course (duration, frequency, number of consecutive night shifts worked, shift length and rest periods) were associated with breast cancer incidence.

## Methods

### Study design and population

We used data from the UK Biobank, a large-scale biomedical database and research resource in the United Kingdom (UK). All individuals aged 40-69 years, registered with the UK’s National Health Service, and residing within 40 km of one of 22 assessment centers in England, Wales, and Scotland, were invited. Over 500,000 participants underwent baseline assessment (2006-2010), yielding a 5.5% participation rate. Individuals were repeatedly invited to complete touchscreen questionnaires, detailed interviews, and to provide physiological measurements, multimodal imaging, and biological samples.

The flow chart (Figure 1) describes exclusion of participants for the current analyses in detail. Men (n = 229,067) and women lost to follow-up (n = 691) were excluded from the full cohort.

**Figure 1:**
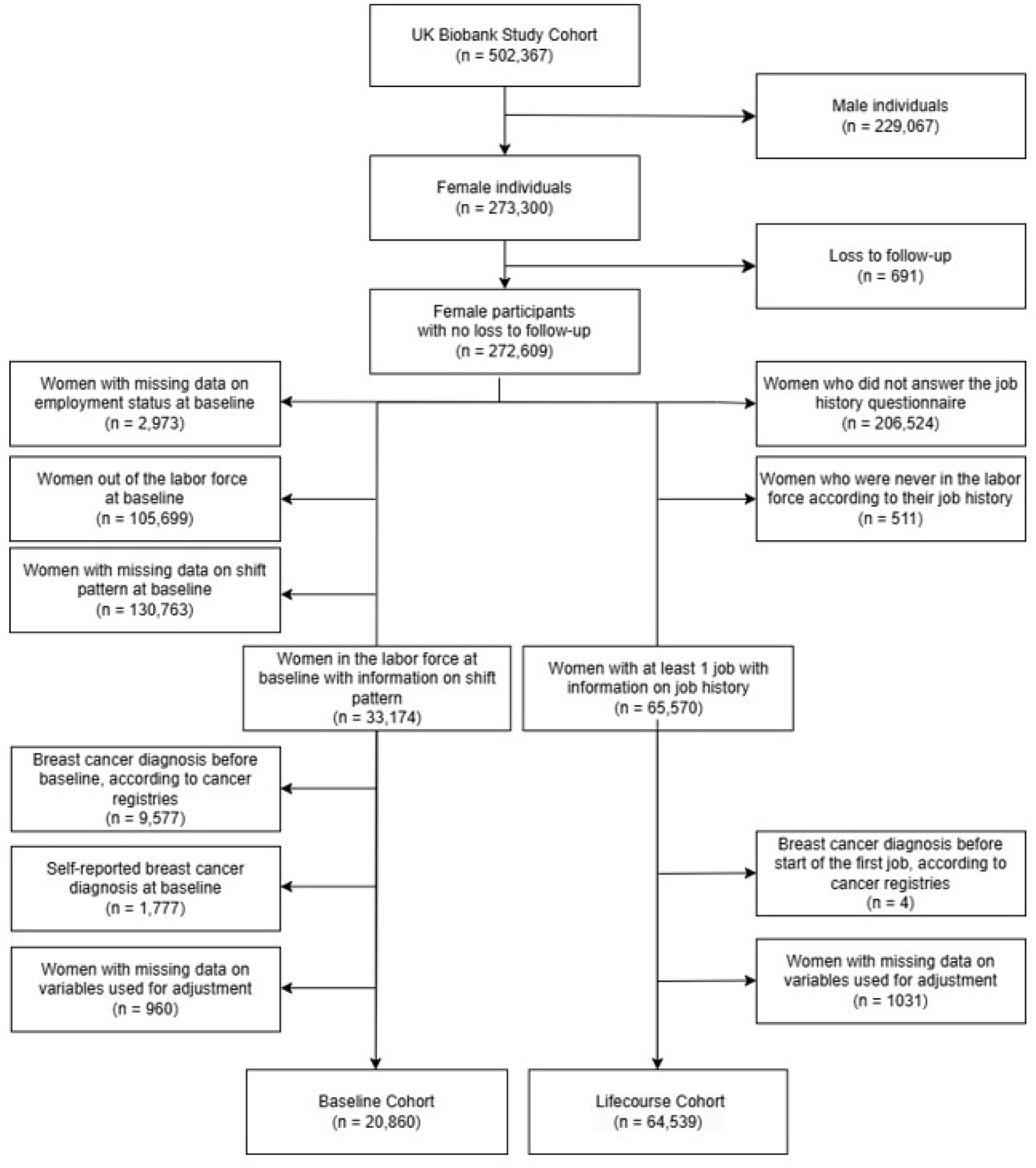
Selection of the participants who were included in the analyses.

To prospectively assess the effect of night shift work and breast cancer incidence (Aim 1) and the moderation effect of sleep patterns and menopausal status (Aims 2 and 3), we used data from a first cohort (Baseline Cohort). This Baseline Cohort consisted of women without a breast cancer diagnosis at baseline (according to cancer registries or based on self-reported diagnosis), who were in the labor force at baseline, with available data on their shift work pattern at baseline. Follow-up for the Baseline Cohort started at baseline and ended on May 13, 2022, the date of the last update of the UK Biobank via linkage with cancer registries, before data extraction.

To assess the effect of various night shift work characteristics and breast cancer incidence (Aim 4), we used data from a second cohort (Lifecourse Cohort) consisting of women with an available job history questionnaire, which detailed the entire working life course, including information on the characteristics of night shift work. Women with a breast cancer diagnosis before starting their first job were excluded. Breast cancer cases were assessed from the reported year of start of the first job in the job history until May 13, 2022.

### Exposure assessment

#### Night shift work

Employment details were collected through a touchscreen questionnaire. For the Baseline Cohort, participants were grouped into day workers or night shift workers based on their response to the baseline question: “Does your main job involve night shifts?”. Night shifts were defined as “a work schedule that involves working through the normal sleeping hours, for instance working through the hours from 12 am to 6 am”. Participants who answered “Never” or “Rarely” were categorized as day workers, while those who answered “Always,” “Usually,” or “Sometimes” were classified as night shift workers.

For the Lifecourse Cohort, we used a job history questionnaire, which documented the entire working life course. For each position held, participants reported the start and end years of employment and specified whether the job involved night shift work (defined in this questionnaire as “at least 3 hours of work between midnight and 5 am”). In this cohort, we categorized participants as either “ever night shift workers” if they reported at least one job involving night shift work in their job history, or “never night shift workers” if they reported none. If night shifts were reported, participants provided additional details, including: the length of a night shift in hours, the number of night shifts worked in a month, the number of nights worked in a row before a rest day, and the number of rest days after a block of night shifts. Participants were instructed to answer based on the most common night shift patterns for each position held. As the questions for each night shift characteristic allowed participants to provide specific numbers or lengths, we recoded them into categories, based on the most common night shift work policies (34–36) and relevant studies on the subject (14,15). This was done to enable comparisons between our results and existing studies.

We computed five life course night shift characteristics: life course duration, monthly frequency, number of consecutive night shifts, length of each night shift, and number of rest days after a block of night shifts. Using the recorded start and end years for night shift jobs, we calculated the total duration of night shift exposure over the participants’ working life course. This life course exposure duration was grouped into three categories: <5 years, 5-10 years and >10 years of exposure. If night shift work periods overlapped, the overlap time in years was calculated and divided by 2, then added to the non-overlapping duration of each period to avoid overestimating the exposure duration. We calculated the average frequency of night shifts per month over the participants’ working life course, accounting for the duration of each job in years. Each job’s duration was used as a weight to ensure an accurate estimation. Based on the calculated average, participants were then categorized into three groups: <10, 10-20 and >20 night shifts per month. The average frequency of night shifts per month was calculated using the following formula:

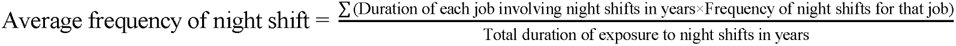

We employed similar weighting methods to determine the length of individual night shifts, the number of consecutive night shifts and number of rest days after a block of night shifts. The length of individual night shifts was grouped into 0–7 hours, 8–12 hours, or >12 hours, reflecting common night shift patterns, particularly 8- and 12-hour shifts. The number of consecutive night shifts was grouped into 1–4 or >4 consecutive nights. To examine recovery time after night shifts, we categorized the number of rest days following a block of night shifts into 3 groups: 1 rest day, 1–3 rest days, and >3 rest days.

#### Sleep

We used self-reported baseline information on sleep traits, including sleep duration and quality, along with information on chronotype.

Sleep duration was assessed with the question: “About how many hours of sleep do you get in every 24 hours? (please include naps)”. We recoded participants’ answers into 2 categories : 7-9 hours, which corresponds to the recommendations (37), and < 7 or > 9 hours, which both are linked to poor health outcomes, including breast cancer, although the evidence regarding the impact of sleeping more than 9 hours on breast cancer remains mixed (38–40).

Sleep quality was assessed based on participants’ answers to four questions: “Do you have trouble falling asleep at night or do you wake up in the middle of the night?”, “How likely are you to doze off or fall asleep during the daytime, when you don’t mean to?”, “How easy do you find getting up in the morning?”, and “Do you have a nap during the day?”. If participants answered yes to at least one of these questions, their sleep quality was considered poor. Otherwise, their sleep quality was considered good.

Chronotype was reported by participants, who selected their type from several options ranging from morning person to evening person. For our analyses, we categorized participants into two categories: ‘morning chronotype’, if participants considered themselves “Definitely a ‘morning’ person” or “More a ‘morning’ than an ‘evening’ person”, and ‘evening chronotype’ if participants reported considering themself “Definitely an ‘evening’ person” or “More an ‘evening’ than a ‘morning’ person”.

#### Menopausal status

Baseline data on menopausal status were obtained from participants in the Baseline Cohort through the question: “Have you had your menopause (periods stopped)?”. Women who answered “Yes” were classified as postmenopausal, those who answered “No” as premenopausal, and women who selected “Prefer not to answer,” “Not sure, had a hysterectomy,” or “Not sure, other reason” were classified as missing.

### Outcome assessment: breast cancer

Incident breast cancer cases were identified through linkage with cancer registries, using the International Classification of Diseases (ICD) codes: ICD-10 (code C50) and ICD-9 (code 174). If a participant received multiple breast cancer diagnoses during her life course, only the first diagnosis was retained for outcome assessment.

### Covariates

Within a causal framework, we selected the set of covariates using a directed acyclic graph (DAG) for all aims (Figure 2 for a simplified DAG and Supplemental Figure 1 for the full DAG). The minimal sufficient adjustment set for estimating the effect of night shift work on breast cancer incidence comprised four variables assessed at baseline: age, ethnicity, education, and parity. Ethnicity was assessed using the question “What is your ethnic group?” and then categorized as White, Asian, Black and Mixed (41). Education was measured based on the question “Which of the following qualifications do you have? (You can select more than one)”, and grouped into College, A levels, O levels/GCSEs, CSEs or equivalent, and Compulsory (42). Parity was assessed with the question “How many children have you given birth to? (Please include live births only)” and categorized as 0, 1, or ≥ 2 children (43). We excluded women with missing data on any of those variables from the analyses (see Figure 1).

**Figure 2:**
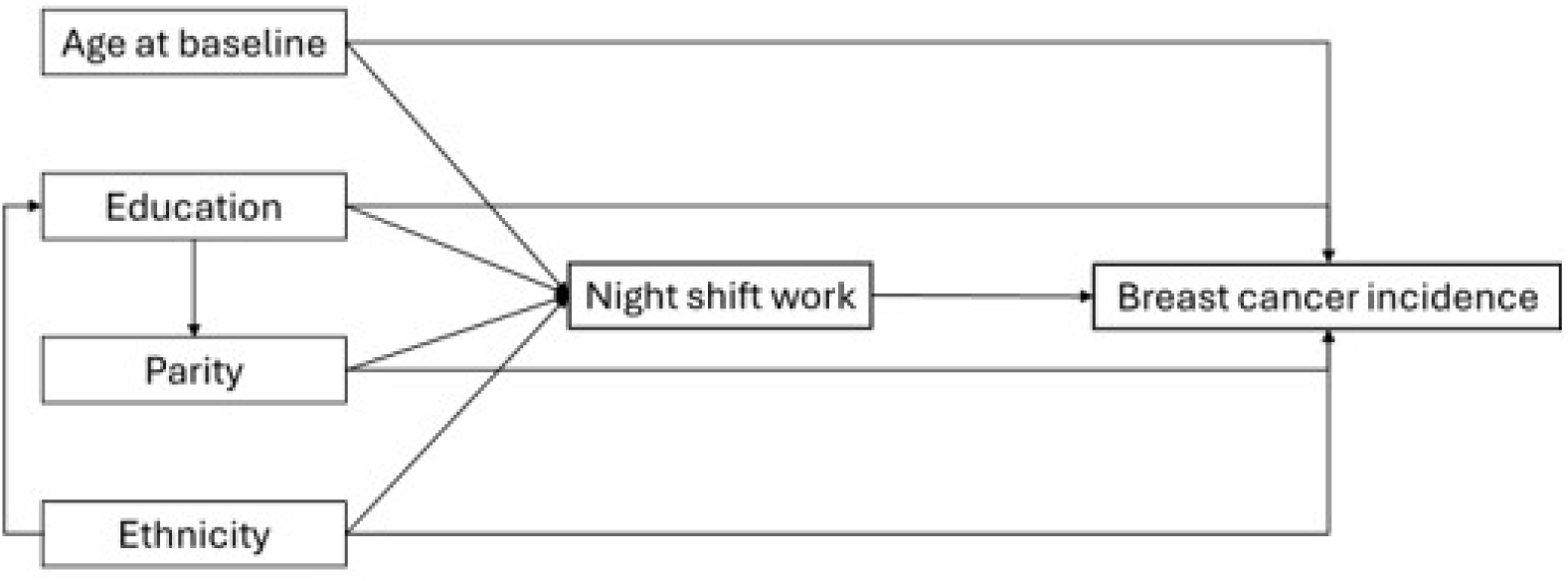
Simplified Directed Acyclic Graph for night shift work and breast cancer incidence.

### Statistical analyses

Baseline characteristics were reported as means and standard deviations for continuous variables, or as counts and percentages for categorical variables.

In the Baseline Cohort, we used Cox proportional hazard regression to estimate multivariable-adjusted hazard ratios (HR) and 95% confidence intervals (95% CIs) to assess the effect of night shift work at baseline on breast cancer incidence (Aim 1). Analyses were performed with adjustment for age at baseline only as well as with adjustment for the final set of covariates.

In the same cohort, stratified analyses explored whether sleep quality, quantity, or chronotype modified the night shift work-breast cancer relationship (Aim 2). We compared sleep duration, quality, and chronotype among night shift workers, allowing us to identify potential subgroups with differing risks of breast cancer related to night shift work. A further stratified analysis assessed whether menopausal status (premenopausal versus postmenopausal) at baseline modified the association between night shift work and breast cancer (Aim 3).

We used the Lifecourse Cohort, to assess the effect of night shift work on breast cancer incidence, and more specifically the effect of various night shift characteristics (durations, frequencies, consecutive nights worked, and rest periods) on breast cancer incidence (Aim 4).

We verified the assumptions for Cox models through visual inspection of residual and statistical tests and no violations were detected. We also performed a sensitivity analysis in the Baseline Cohort by reclassifying participants who answered “Sometimes” as day workers instead of night shift workers.

R version 4.3.1 (2023-06-16 ucrt) - ‘Beagle Scouts’, was used to conduct the analyses.

## Results

### Participant Characteristics

The Baseline Cohort included 20,680 breast cancer-free women who were in the labor force at baseline, with data on shift patterns and covariates. The Lifecourse Cohort comprised 64,539 women who were breast cancer-free before starting their first job, completed a detailed job history questionnaire with at least one period of employment in the labor force, and with data on all covariates.

Table 1 presents the descriptive characteristics of the cohorts at baseline. All detailed characteristics corresponding to the variables used for the creation of the full DAG, with their distribution across the two cohorts, are presented in Supplemental Table 1. For the Baseline Cohort, the mean age at baseline was 51.7 years old. For the Lifecourse Cohort, the mean age at baseline was 55.6 years old. Most participants were of white ethnicity, had attended college, and had given birth to two or more children.

**Table 1:**
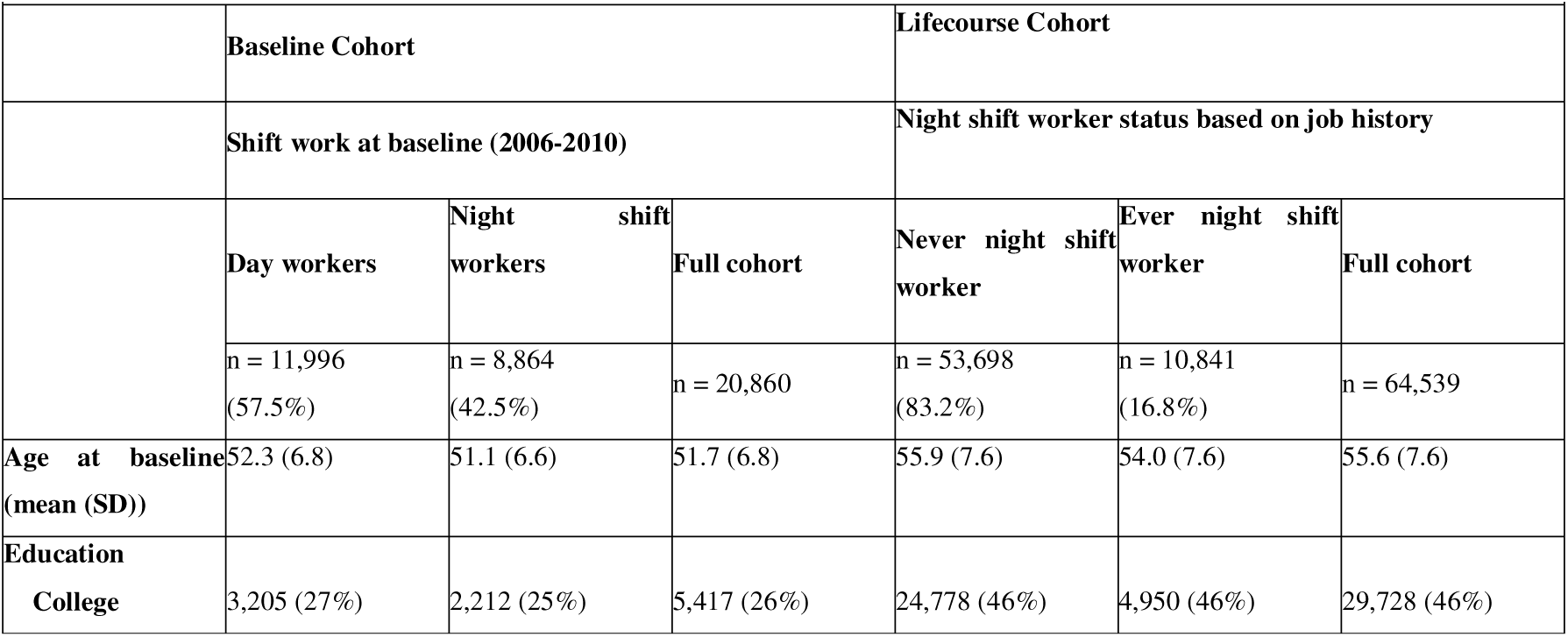
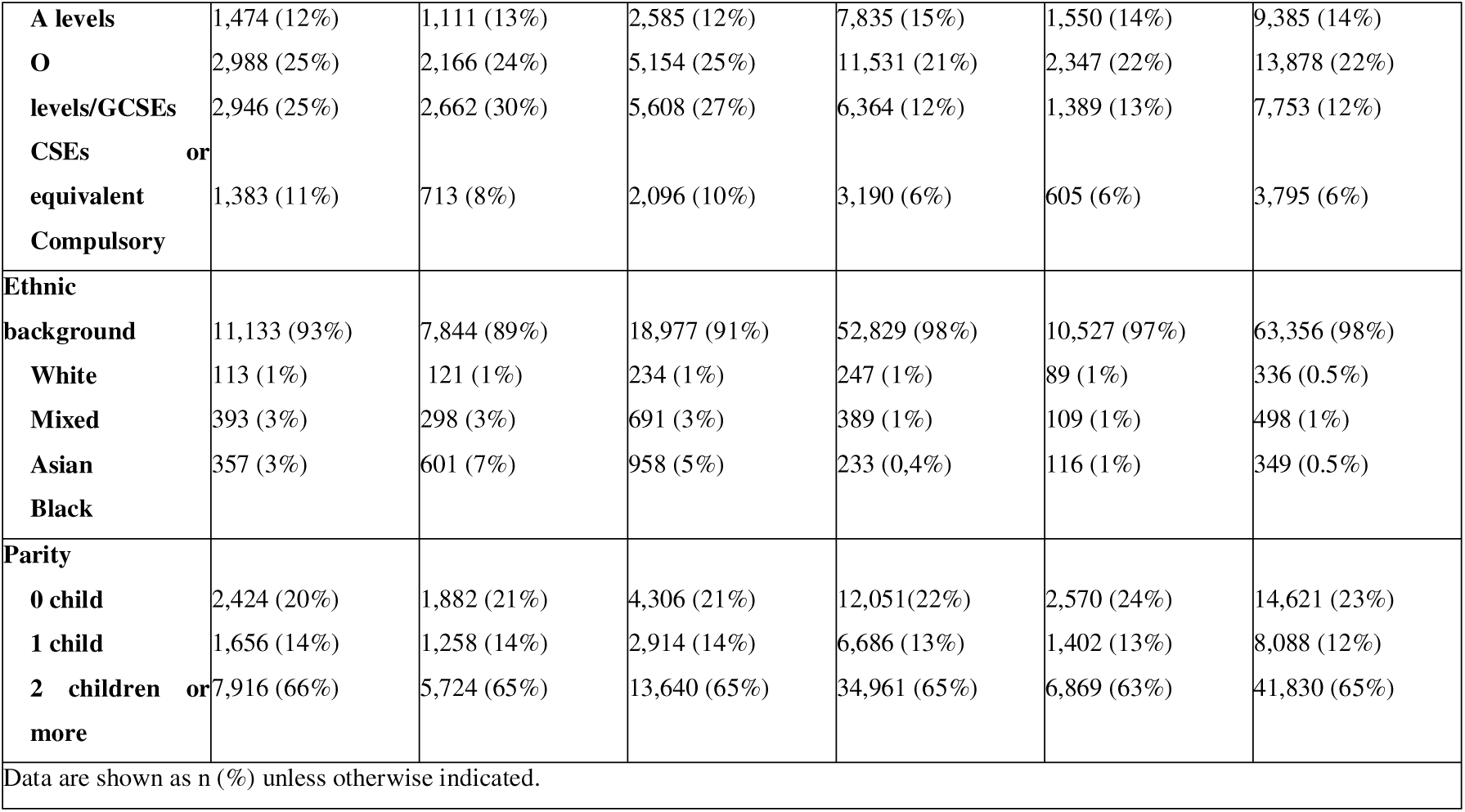
Participant Characteristics at Baseline: Baseline Cohort and Lifecourse Cohort.

The participants of the Lifecourse Cohort had between 1 and 20 different work periods throughout their careers, with an average of 4 distinct jobs per participant over their working life course. These work periods began between 1949 and 2015. In the ever night shift worker subgroup, participants held between 1 and 11 jobs involving night shifts. The median duration of night shift work was 10 years (first quartile: 4 years; third quartile: 27 years). Additionally, 100 participants had overlapping night shift work periods during their careers.

### Night shift work and breast cancer incidence

In the Baseline Cohort, the median follow-up duration was 14 years, during which 688 breast cancer cases were diagnosed, resulting in an incidence rate of 236 cases per 100,000 person-years. The adjusted HR for breast cancer incidence in relation to being a night shift worker at baseline was 0.95 (95% CI: 0.85-1.06) (Table 2). The sensitivity analysis, in which participants who answered ‘Sometimes’ were reclassified as day workers instead of night shift workers, did not alter the results.

**Table 2:**
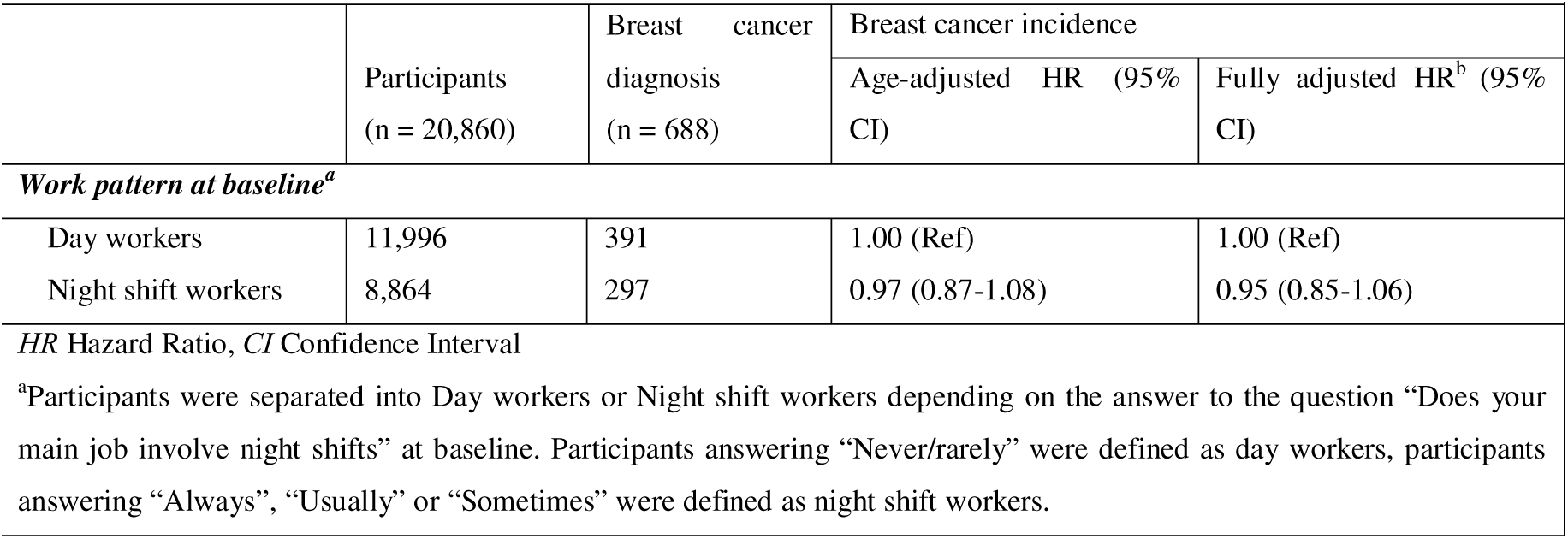
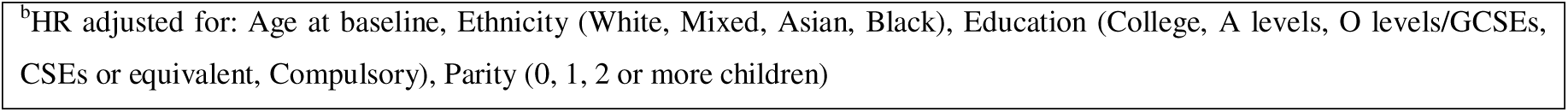
Cox regression hazard ratios for shift work pattern at baseline^a^ with breast cancer, Baseline Cohort.

|  |  |  |  |  |
| --- | --- | --- | --- | --- |
|  | Participants<br>(n = 20,860) | Breast cancer<br>diagnosis<br>(n = 688) | Breast cancer incidence |  |
|  |  |  | Age-adjusted HR (95% CI) | Fully adjusted HR<sup>b</sup> (95% CI) |
| Work pattern at baseline<sup>a</sup> |  |  |  |  |
| Day workers | 11,996 | 391 | 1.00 (Ref) | 1.00 (Ref) |
| Night shift workers | 8,864 | 297 | 0.97 (0.87-1.08) | 0.95 (0.85-1.06) |
| HR Hazard Ratio, CI Confidence Interval |  |  |  |  |
| <sup>a</sup>Participants were separated into Day workers or Night shift workers depending on the answer to the question “Does your main job involve night shifts” at baseline. Participants answering “Never/rarely” were defined as day workers, participants answering “Always”, “Usually” or “Sometimes” were defined as night shift workers. |  |  |  |  |

<sup>b</sup>HR adjusted for: Age at baseline, Ethnicity (White, Mixed, Asian, Black), Education (College, A levels, O levels/GCSEs, CSEs or equivalent, Compulsory), Parity (0, 1, 2 or more children)

In the Lifecourse Cohort, the median follow-up duration was 50 years, during which 4,455 cases of breast cancer were diagnosed, corresponding to an incidence rate of 138 cases per 100,000 person-years. The adjusted HR for breast cancer incidence in relation to being a night shift worker over the working life course was 1.04 (95% CI: 0.96-1.13) (Table 3).

**Table 3:** Cox regression hazard ratios for night shift worker status^a^ and by night shift work characteristics, Lifecourse Cohort.

|  |  |  |  |  |
| --- | --- | --- | --- | --- |
|  | Participants<br>(n = 64,539) | Breast cancer<br>diagnosis<br>(n = 4,455) | Breast cancer incidence |  |
|  |  |  | Age-adjusted<br>HR (95% CI) | Fully adjusted<br>HR<sup>b</sup> (95% CI) |
| ***Night shift worker status according to job history<sup>a</sup>*** |  |  |  |  |
| Never night shift worker | 53,698 (83%) | 3,780 (7%) | 1.00 (Ref) | 1.00 (Ref) |
| Ever night shift worker | 10,841 (17%) | 675 (6%) | 1.03 (0.95-1.12) | 1.04 (0.96-1.13) |
| ***Night shift work characteristics according to job history*** |  |  |  |  |
| Life course duration of night shift work in years |  |  |  |  |
| Never night shift worker | 53,698 (83%) | 3,780 (7%) | 1.00 (Ref) | 1.00 (Ref) |
| Less than 1 year to 5 years | 3,072 (5%) | 194 (6%) | 1.00 (0.86-1.15) | 0.98 (0.84-1.14) |
| 5-10 years | 1,910 (3%) | 114 (6%) | 1.09 (0.91-1.32) | 1.10 (0.91-1.32) |
| >10 years | 5,859 (9%) | 367 (6%) | 1.02 (0.92-1.14) | 1.06 (0.95-1.18) |
| Mean monthly frequency of night shifts |  |  |  |  |
| Never night shift worker | 53,698 (83%) | 3,780 (7%) | 1.00 (Ref) | 1.00 (Ref) |
| < 10 | 7,744 (12%) | 444 (6%) | 1.06 (0.96-1.17) | 1.07 (0.97-1.19) |
| 10-20 | 2,868 (4%) | 210 (7%) | 0.99 (0.86-1.13) | 1.01 (0.88-1.16) |
| >20 | 229 (0.4%) | 21 (9%) | 0.87 (0.57-1.34) | 0.84 (0.55-1.29) |
| Mean length of each night shift in hours |  |  |  |  |
| Never night shift worker | 53,698 (83%) | 3,780 (7%) | 1.00 (Ref) | 1.00 (Ref) |
| 0-7 | 833 (1%) | 57 (7%) | 1.16 (0.89-1.51) | 1.25 (0.96-1.63) |
| 8-12 | 9,205 (14%) | 562 (6%) | 1.02 (0.93-1.11) | 1.03 (0.95-1.13) |
| >12 | 803 (1%) | 56 (7%) | 1.02 (0.78-1.33) | 0.97 (0.75-1.27) |
| Mean number of consecutive night shifts |  |  |  |  |
| Never night shift worker | 53,698 (83%) | 3,780 (7%) | 1.00 (Ref) | 1.00 (Ref) |
| 1-4 | 6,894 (11%) | 395 (6%) | 1.01 (0.91-1.12) | 1.03 (0.92-1.14) |
| >4 | 3,947 (6%) | 280 (7%) | 1.06 (0.94-1.20) | 1.07 (0.95-1.20) |
| Mean number of rest days after block of night shift |  |  |  |  |
| Never night shift worker | 53,698 (83%) | 3,780 (7%) | 1.00 (Ref) | 1.00 (Ref) |
| 1 | 2,825 (4%) | 168 (6%) | 1.04 (0.89-1.21) | 1.03 (0.88-1.20) |
| 1-3 | 4,959 (8%) | 321 (7%) | 1.03 (0.92-1.15) | 1.06 (0.94-1.18) |
| >3 | 3,057 (5%) | 186 (6%) | 1.02 (0.88-1.18) | 1.04 (0.90-1.20) |
| *HR* Hazard Ratio, *CI* Confidence Interval |  |  |  |  |
| Data are n (%) unless indicated. |  |  |  |  |
| <sup>a</sup>Night shift worker status: Participants were considered “ever night shift worker” if they reported at least 1 job involving night shift in their employment history |  |  |  |  |
| <sup>b</sup>HR adjusted for: Age at baseline, Ethnicity (White, Mixed, Asian, Black), Education (College, A levels, O levels/GCSEs, CSEs or equivalent, Compulsory), Parity (0, 1, 2 or more children) |  |  |  |  |

The duration of exposure to night shift work (in years) over a woman’s working life course was not associated with an increased incidence of breast cancer. Specifically, compared to women who never worked in jobs involving night shifts, the total duration of night shift work was not associated with breast cancer incidence for those with less than 5 years in jobs involving night shifts (HR 0.98; 95% CI: 0.84–1.14), 5 to 10 years (HR 1.10; 95% CI: 0.91–1.32), nor more than 10 years (HR 1.06; 95% CI: 0.95–1.18).

The mean monthly frequency of night shift work was not associated with breast cancer incidence, with fully adjusted HRs of 1.07 (95% CI: 0.97-1.19) for women working less than 10 nights a month, 1.01 (95% CI: 0.88-1.16) for women working between 10 and 20 nights a month, and 0.84 (95% CI: 0.55-1.29) for women working more than 20 nights a month.

The length of a typical night shift showed no effect on breast cancer incidence, with fully adjusted HRs of 1.25 (95% CI: 0.96-1.63) for women working night shifts lasting <7 hours, 1.03 (95% CI: 0.95-1.13) for women working night shifts lasting between 7 and 12 hours, and 0.97 (95% CI: 0.75-1.27) for women working night shifts lasting more than 12 hours.

The mean number of consecutive night shifts worked also showed no effect on breast cancer incidence, with fully adjusted HRs of 1.03 (95% CI: 0.92-1.14) for women working between 1 to 4 nights in a row, and 1.07 (95% CI: 0.95-1.20) for women working more than 4 nights in a row.

The number of rest days following a block of night shifts was not related to the risk of developing breast cancer, with fully adjusted HRs of 1.03 (95% CI: 0.88-1.20) for women getting 1 rest day after a block of night shifts, 1.06 (95% CI: 0.94-1.18) for women getting 2 to 3 rest days after a block of night shifts, and 1.04 (95% CI: 0.90-1.20) for women getting more than 3 rest days after a block of night shifts.

### Moderation of sleep, chronotype and menopausal status

Using the Baseline Cohort, we stratified the risk of breast cancer in relation to night shift worker status at baseline by sleep duration, sleep quality, and chronotype. In this cohort, 18,977 women had available data on sleep duration, sleep quality and chronotype. No associations or interactions were found for any of these factors (Table 4). Further analysis by menopausal status at baseline also revealed no associations or interactions on the relationship between night shift work and breast cancer incidence.

**Table 4.** Incidence of breast cancer associated with shift work pattern at baseline^a^, stratified by sleep patterns and menopause, Baseline Cohort.

|  | Participants | Breast cancer<br>diagnosis | Breast cancer incidence |  |
| --- | --- | --- | --- | --- |
|  |  |  | Age-adjusted HR (95% CI) | Fully adjusted HR<sup>b</sup> (95% CI) |
| ***Work pattern at baseline<sup>a</sup> (participants with available data on sleep duration, sleep quality and chronotype only)*** |  |  |  |  |
| Day workers | 10,968 (58%) | 360 (3%) | 1.00 (Ref) | 1.00 (Ref) |
| Night shift workers | 8,009 (42%) | 268 (3%) | 0.95 (0.85-1.06) | 0.95 (0.85-1.06) |
| Total | 18,977 | 628 |  |  |
| ***Sleep duration at baseline<sup>c</sup>*** |  |  |  |  |
| Sleep 7-9 hours /24 hours |  |  |  |  |
| Day workers | 7,804 (41%) | 258 (3%) | 1.00 (Ref) | 1.00 (Ref) |
| Night shift workers | 5,260 (28%) | 178 (3%) | 0.90 (0.79-1.04) | 0.91 (0.79-1.04) |
| Total | 13,064 (69%) | 436 (3%) |  |  |
| Sleep <7 or >9 hours/24 hours |  |  |  |  |
| Day workers | 3,164 (17%) | 102 (3%) | 1.00 (Ref) | 1.00 (Ref) |
| Night shift workers | 2,749 (15%) | 90 (3%) | 1.07 (0.87-1.32) | 1.06 (0.86-1.32) |
| Total | 5,913 (31%) | 192 (3%) |  |  |
| ***Sleep quality at baseline<sup>d</sup>*** |  |  |  |  |
| Good sleep quality |  |  |  |  |
| Day workers | 1,096 (6%) | 30 (3%) | 1.00 (Ref) | 1.00 (Ref) |
| Night shift workers | 715 (4%) | 27 (4%) | 1.28 (0.88-1.90) | 1.35 (0.87-2.09) |
| Total | 1,811(10%) | 57 (3%) |  |  |
| Poor sleep quality |  |  |  |  |
| Day workers | 9,872 (52%) | 330 (3%) | 1.00 (Ref) | 1.00 (Ref) |
| Night shift workers | 7,294 (38%) | 241 (3%) | 0.92 (0.82-1.04) | 0.92 (0.82-1.04) |
| Total | 17,166 (90%) | 571 (3%) |  |  |
| ***Chronotype at baseline<sup>e</sup>*** |  |  |  |  |
| Morning chronotype |  |  |  |  |
| Day workers | 6,768 (36%) | 221 (3%) | 1.00 (Ref) | 1.00 (Ref) |
| Night shift workers | 4,380 (23%) | 123 (3%) | 0.96 (0.82-1.12) | 0.93 (0.79-1.10) |
| Total | 11,148 (59%) | 344 (3%) |  |  |
| Evening chronotype |  |  |  |  |
| Day workers | 4,200 (22%) | 139 (3%) | 1.00 (Ref) | 1.00 (Ref) |
| Night shift workers | 3,629 (19%) | 145 (4%) | 0.90 (0.76-1.06) | 0.88 (0.74-1.04) |
| Total | 7,829 (41%) | 284 (4%) |  |  |
| ***Work pattern at baseline<sup>a</sup> (participants with available data on menopausal status at baseline only)*** |  |  |  |  |
| Day workers | 9,912 (58%) | 321 (3%) | 1.00 (Ref) | 1.00 (Ref) |
| Night shift workers | 7,321 (42%) | 242 (3%) | 0.98 (0.87-1.10) | 0.98 (0.87-1.10) |
| Total | 17,233 | 563 |  |  |
| **Menopausal status at baseline<sup>f</sup>** |  |  |  |  |
| Premenopausal |  |  |  |  |
| Day workers | 4,267 (25%) | 131 (3%) | 1.00 (Ref) | 1.00 (Ref) |
| Night shift workers | 3,565 (21%) | 109 (3%) | 0.96 (0.80-1.15) | 0.98 (0.81-1.18) |
| Total | 7,832 (45%) | 240 (3%) |  |  |
| Postmenopausal |  |  |  |  |
| Day workers | 5,645 (33%) | 190 (3%) | 1.00 (Ref) | 1.00 (Ref) |
| Night shift workers | 3,756 (22%) | 133 (4%) | 0.99 (0.85-1.16) | 0.95 (0.81-1.11) |
| Total | 9,401 (55%) | 323 (3%) |  |  |
| *HR Hazard Ratio, CI Confidence Interval*<br>Data are n (%) unless indicated.<br><sup>a</sup>Participants were separated into Day workers or Night shift workers depending on the answer to the question “Does your main job involve night shifts” at baseline. Participants answering “Never/rarely” were defined as day workers, participants answering “Always”, “Usually” or “Sometimes” were defined as night shift workers.<br><sup>b</sup>HR adjusted for: Age at baseline, Ethnicity (White, Mixed, Asian, Black), Education (College, A levels, O levels/GCSEs, CSEs or equivalent, Compulsory), Number of live births (0, 1, 2 or more)<br><sup>c</sup>Sleep duration: "About how many hours sleep do you get in every 24 hours? (please include naps)"<br><sup>d</sup>Assessment of sleep quality was based on the questions: “Do you have trouble falling asleep at night or do you wake up in the middle of the night?”, “How likely are you to doze off or fall asleep during the daytime, when you don’t mean to?”, “How easy do you find getting up in the morning?”, and “Do you have a nap during the day?” If participants reported having trouble falling asleep, staying awake during the day, getting up, or needing to nap, their sleep quality was considered poor.<br><sup>e</sup>Chronotype: morning chronotype if the participant reported considering themselves “Definitely a ‘morning’ person” or “More a ‘morning’ than an ‘evening’ person”, and evening type if the participant reported considering themselves “Definitely a ‘evening’ person” or “More a ‘evening’ than an ‘morning’ person”.<br><sup>f</sup>Menopausal status: "Have you had your menopause (periods stopped)?" (yes/no) |  |  |  |  |

## Discussion

Our large prospective study provides robust evidence on the effect of night shift work on breast cancer incidence, exploring life course characteristics of night shift work while accounting for sleep habits, chronotype, and menopausal status. Specifically, we examined (1) whether night shift work was associated with breast cancer incidence, whether this association was modified by (2) sleep characteristics and (3) menopausal status, and (4) whether detailed working life course characteristics of night shift work (such as duration, frequency, number of consecutive night shifts worked, shift length, and rest periods) were linked to breast cancer incidence.

### Night shift work and breast cancer

We found no effect of night shift work on breast cancer incidence. Our findings align with previous cohort studies (27,44), including the 2016 UK Biobank study by Travis et al., which also reported no association (26). However, our study improves on prior UK Biobank analyses by using updated follow-up data (through 2022) and on our use of a DAG to identify confounders for minimal adjustment.

In contrast, the positive association observed in case–control studies (12,15,24,25,45) may be misleading due to a higher risk of bias inherent in their design. Several meta-analyses (46–49) have reinforced the null findings from cohort studies. Taken together, the consistency of null findings across multiple well-designed cohort studies, including ours with relatively long follow-up time, strongly suggests that night shift work does not have an effect on breast cancer incidence.

### Sleep and chronotype

We found no evidence that sleep duration, sleep quality and chronotype modifies the association between night shift work and breast cancer. This aligns with the 2016 UK Biobank study by Travis et al., which reported no impact of the number of hours of sleep and the diurnal preference on breast cancer incidence (26). Similarly, the IARC’s 2019 evaluation concluded that there is no evidence regarding the interaction of diurnal preference with the association between night shift work and breast cancer (16).

The classification of participants into good or poor sleep quality merits caution. Indeed, in our study, nap-taking was used as a proxy for poor sleep quality, but it may also reflect sleep fragmentation – a common occurrence after night shifts – rather than restorative naps, which could be part of the natural adaptation to disrupted sleep cycles following night shifts.

### Menopause

Some studies suggested that postmenopausal women could have a lower incidence of breast cancer compared with premenopausal women when working night shifts (6,14,15). This could be explained considering that premenopausal women could experience more estrogen dysregulation due to circadian rhythm disruption (7,20,21). In our study, we did not find a modifying effect of menopausal status and a null association was observed both in premenopausal and postmenopausal women.

### Night shift work across the entire working life course and breast cancer

Although we found no association between night shift work and breast cancer incidence, current evidence remains limited due to a lack of detailed assessments of night shift work characteristics across the entire working life course. Addressing this gap, our study used a bi-directional design – which combines a prospective approach (Baseline Cohort) with retrospective data collection (Lifecourse Cohort) – to provide a more comprehensive assessment of night shift work across the entire working life course. Our results showed that key characteristics of night shift work over the working life course – including exposure duration, monthly frequency, shift length, number of consecutive night shifts, and rest days after night shift blocks – were not associated with an increased incidence of breast cancer.

To our knowledge, this is the first study to evaluate such a comprehensive range of night shift work characteristics, particularly rest days and consecutive night shifts, while accounting for exposure across the entire working life course. Unlike traditional carcinogens, where risk is typically linked to cumulative exposure duration and dose-response relationships, night shift work over the life course does not appear to follow this type of cumulative risk mechanism.

Several studies have suggested an increased risk of breast cancer among women with long-term exposure to night shift work. For example, two meta-analyses based on both cohort and case-control studies found a positive association between long-term night shift work and breast cancer incidence (with statistically significant increases observed after 10, 20, and 30 years of exposure) (14,22). Some studies and meta-analyses even reported a dose–response relationship, with breast cancer incidence increasing with the number of years of night shift work (21,22,50,51). Our results contradict these findings and are more consistent with those of the meta-analysis by Travis et al. (2016), which included the first UK Biobank study on this topic and reported no association between long-term night shift work (>20 years) and breast cancer incidence (26), and those of a recent dose-response meta-analysis (17). One possible explanation lies in differences in exposure assessment. In our study, night shift work was quantified as a cumulative duration based on the participant’s entire job history, whereas many other studies have focused on night shift work during the follow-up period only. Our approach provides a more realistic estimate of lifetime exposure. However, it assumes a linear dose– response relationship and may overlook the existence of sensitive periods or threshold effects, such as heightened vulnerability to circadian disruption during specific life stages.

Regarding frequency of night shift work, a meta-analysis conducted by Hong et al. found that high-frequency night shift work (>5 shifts/week) is a risk factor to develop breast cancer (14), which aligns with the results of the 2018 pooled case-control study of Cordina-Duverger et al. finding a positive association for premenopausal women working at least 3 night shifts per week (15). Notably, these studies assessed night shift work on a weekly basis, whereas our study used a weighted monthly average frequency, calculated from the entire job history of each participant. This offers a more accurate measure of long-term exposure and its potential cumulative impact. Since cancer is a disease with a long latency period, it is essential to capture prolonged and repeated exposures over time.

Finally, the analysis of consecutive night shifts and rest periods between shifts revealed no association with breast cancer incidence. These findings do not support the hypothesis that insufficient recovery time exacerbates breast cancer incidence. It is important to note that in recent years, especially during the follow-up of the Lifecourse Cohort, work schedule policies has evolved to limit consecutive night shifts, allowing for more rest days between shifts, for example due to the 1998 Working Time Regulations or the 2003 European Parliament Directive (36,52,53). However, our study was not designed to evaluate the impact of these policies. Further studies considering contemporary policies about night shift work are needed to evaluate their impact and if further adjustments are necessary.

### Strengths and limitations

A key strength lies in the complementary design of the two cohorts. The Baseline Cohort provided data on night shift work that minimized recall bias, while the Lifecourse Cohort enabled analyses of detailed night shift work characteristics across the entire working life course. Combining these approaches enhanced the reliability of our findings. Both cohorts had a relatively long follow-up and had large sample sizes, with sufficient cases for analyzing the main effects of night shift work. In both cohorts, the exposure chronologically preceded the outcome, and the outcome was based on registry data. Additionally, detailed data on other potential risk factors and the use of a DAG strengthened our methodology by clarifying the minimal sufficient adjustment set of confounding factors.

Despite these strengths, our study has several limitations. First, it relies on self-reported data for exposure assessment and covariates, which may introduce biases. Night shift work definition differed between our two analytical cohorts, potentially contributing to heterogeneity in findings, a known issue in prior studies (10). In the Lifecourse Cohort, job histories were based on retrospective self-reports, requiring participants to recall detailed work characteristics over their entire working life course, potentially leading to recall bias. Self-reported data on sleep patterns and chronotype may have introduced measurement errors, and we did not account for changes in sleep patterns and menopausal status during follow-up, potentially leading to misclassification.

Second, it has been shown that the UK Biobank population is not representative of the general UK population (54). Therefore, selection bias may affect our results and compromise their external validity (55). Furthermore, we applied strict exclusion criteria due to the lack of data on night shift work, which led to a reduced sample size in our analytical cohorts compared to the original dataset. This may have introduced an additional selection bias and limited the generalizability of our results.

Third, survivor bias is inherent to the UK biobank’s design, which recruited participants aged 40-69 years. This may limit the generalizability of our findings to populations who survived to the study period. However, the survivor bias may likely be limited, as the probability of dying from cancer before the age of 40 is low (56). On the other hand, the average follow-up of 14 years may have been insufficient to capture the development of all breast cancers in the sample.

Fourth, in the UK Biobank, information on whether participants emigrated or withdrew from the study was only available until 2017. We estimated that censoring may not have been accurately recorded beyond this date for approximately 470 individuals in our study. Given the small number of affected participants, any resulting bias is likely minimal.

Fifth, complete cancer incidence data for participants were not available until the end of follow-up: complete data from Wales were available only up to 2017, from England up to 2021, and from Scotland up to 2022 (57). This incomplete follow-up may have led to some underestimation of breast cancer cases in this subgroup, potentially affecting the accuracy of our findings.

Finally, our study also did not distinguish between consistent and intermittent night shift workers, nor did it address factors like workplace light exposure or other environmental exposures. Future research should incorporate these variables for a more comprehensive understanding.

### Future perspectives

Our findings alleviate concerns about the overall risks of night shift work for breast cancer incidence. However, night shift work is associated with poor health behaviors and chronic stress, both recognized as carcinogenic risk factors (58). Additionally, night shift workers often face socio-economic disparities and limited access to healthcare (59). Therefore, it is crucial to maintain working time regulations, optimize shift schedules, and provide resources to help workers manage the health risks linked to disrupted circadian rhythms. In addition, future research should integrate genetic, hormonal, and lifestyle factors, along with objective and long-term measures of light exposure and circadian markers, to clarify the biological mechanisms linking night shift work and breast cancer as suggested by the IARC (16) and the US National Toxicology Program (60).

## Conclusion

In conclusion, this study indicates that there is no effect of night shift work on breast cancer in the examined population. This adds to the current knowledge base on night shift work and breast cancer, suggesting the relationship may be more nuanced than previously thought. The lack of a direct association highlights the importance of exploring individual variability in response to night shift work. Future research should continue to address these complexities to provide clearer guidance for public health interventions and workplace policies.

## Additional information

## Acknowledgements

This research has been conducted using the UK Biobank Resource under application number 86247.

## Authors’ contributions

JS, BWAvdL and SC designed the study. JS and BWAvdL conducted the analyses, and JS cleaned the data. JS, BWAvdL and SC interpreted the data and produced the figures. JS, BWAvdL and SC drafted the first manuscript. All authors critically revised the manuscript, approved the final version, and agreed to be accountable for all aspects of the work to ensure the accuracy and integrity of any part of the study. The corresponding author (BWAvdL) confirms full access to all data in the study and final responsibility for the decision to submit the manuscript for publication.

## Ethics approval and consent to participate

All UK Biobank’s participants provided written consent, and the study protocol was approved by the North West Multicenter Research Ethics Committee in the United Kingdom.

## Data availability

The UK Biobank is an open access resource, available at https://www.ukbiobank.ac.uk/researchers/.

## Competing interests

The authors declare no conflict of interest.

## Funding information

The author(s) received no specific funding for this work.

## Notes

### Competing Interest Statement

The authors have declared no competing interest.

### Author Declarations

The UK Biobank is an open access resource, available at https://www.ukbiobank.ac.uk/researchers/.

