## Supplemental Figure 1 for "Night shift work and breast cancer incidence among women: a population-based cohort study"

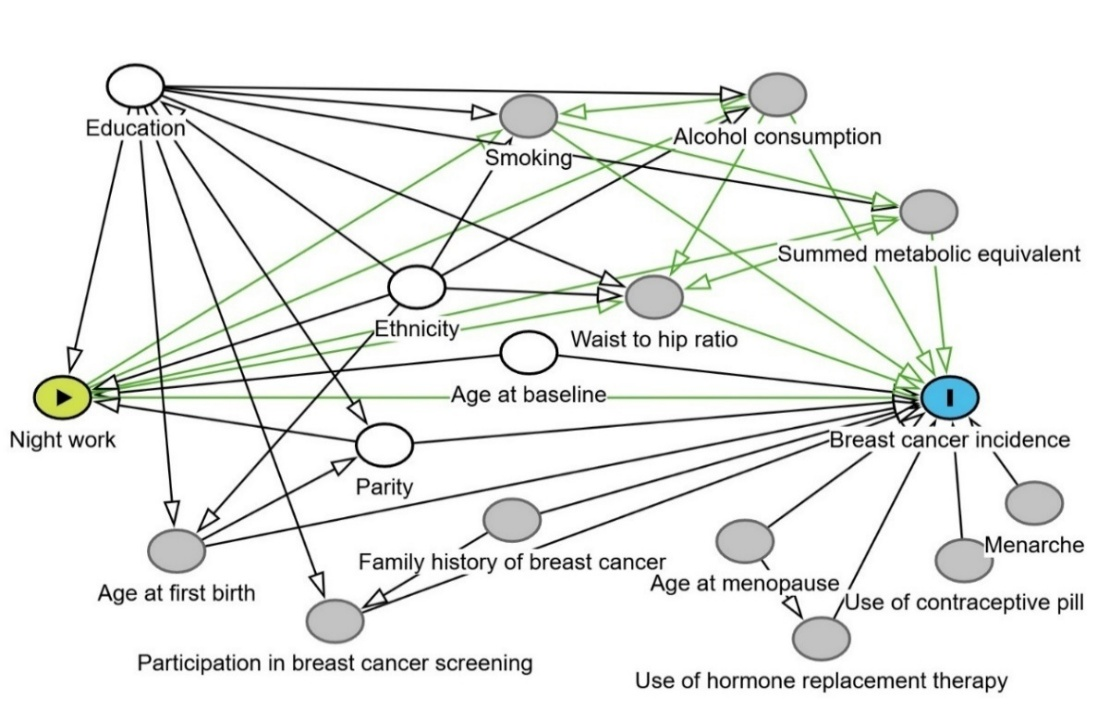
**Supplemental figure 1**: Full Directed Acyclic Graph for night shift work and breast cancer incidence
