## Supplemental Table 1 for "Night shift work and breast cancer incidence among women: a population-based cohort study"

| **Supplemental table 1:** Participant Characteristics at Baseline: Comparison by Baseline Shift Patterns (Baseline Cohort) and Night Worker Status (Lifecourse Cohort) | | | | | | | | | |
| --- | --- | --- | --- | --- | --- | --- | --- | --- | --- |
|  | **Baseline Cohort** | | | **Lifecourse Cohort** | | | | | |
|  | **Shift work at baseline (2006-2010)** | | | **Night shift worker status based on job history** | | | | | |
|  | **Day workers** | **Night shift workers** | **Full cohort** | **Never night shift worker** | | **Ever night shift worker** | | **Full cohort** | |
|  | N = 11,996  (57.5%) | N = 8,864  (42.5%) | N = 20,860 | N = 53,698  (83.2%) | | N = 10,841  (16.8%) | | N = 64,539 | |
| **Age at baseline (mean (SD))** | 52.3 (6.8) | 51.1 (6.6) | 51.7 (6.8) | 55.9 (7.6) | | 54.0 (7.6) | | 55.6 (7.6) | |
| **Weight in kg in 2014 (mean (SD))**  **Missing** | 72.6 (15.0)    50 (1%) | 74.0 (15.6)    40 (0.4%) | 73.2 (15.3)    90 (0.4%) | 70.1 (13.2)    88 (0.2%) | | 72.1 (14.6)    19 (0.2%) | | 70.5 (13.5)    107 (0.2%) | |
| **Height in cm in 2014 (mean (SD))**  **Missing** | 162.8 (6.3)    40 (0.3%) | 162.8 (6.4)    34 (0.4%) | 162.8 (6.3)    74 (0.4%) | | 163.4 (6.1)    62 (0.1%) | | 163.9 (6.2)    9 (0.1) | | 163.5 (6.1)    71 (0.1%) |
| **BMI at recruitment (mean (SD))**  **Missing** | 27.4 (5.4)    50 (0.4%) | 27.9 (5.7)    44 (0.5%) | 27.6 (5.5)    94 (0.5%) | | 26.3 (4.8)    98 (0.2%) | | 26.8 (5.3)    20 (0.2%) | | 26.4 (4.9)    118 (0.1%) |
| **Waist to hip ratio at recruitment (mean (SD))**  **Missing** | 0.82 (0.07)  43 (0.4%) | 0.82 (0.07)  30 (0.3%) | 0.82 (0.07)  73 (0.3%) | | 0.81 (0.07)  65 (0.1%) | | 0.81 (0.07)  12 (0.1%) | | 0.81 (0.07)  77 (0.1%) |
| **MET at recruitment**  **< 500**  **500-5000**  **> 5000**  **Unknown* or missing** | 1,177 (10%)  6,023 (50%)  1,950 (16%)  2,846 (24%) | 737 (8%)  4,183 (47%)  1,643 (19%)  2,301 (26%) | 1,914 (9%)  10,206 (49%)  3,593 (17%)  5,147 (25%) | | 6,643 (12%)  33,202 (62%)  4,684 (9%)  9,169 (17%) | | 1,211 (11%)  6,584 (61%)  1,299 (12%)  1,747 (16%) | | 7,854 (12%)  39,786 (62%)  5,983 (9%)  10,916 (17%) |
| **Education**  **College**  **A levels**  **O levels/GCSEs**  **CSEs or equivalent**  **Compulsory** | 3,205 (27%)  1,474 (12%)  2,988 (25%)  2,946 (25%)  1,383 (11%) | 2,212 (25%)  1,111 (13%)  2,166 (24%)  2,662 (30%)  713 (8%) | 5,417 (26%)  2,585 (12%)  5,154 (25%)  5,608 (27%)  2,096 (10%) | | 24,778 (46%)  7,835 (15%)  11,531 (21%)  6,364 (12%)  3,190 (6%) | | 4,950 (46%)  1,550 (14%)  2,347 (22%)  1,389 (13%)  605 (6%) | | 29,728 (46%)  9,385 (14%)  13,878 (22%)  7,753 (12%)  3,795 (6%) |
| **Ethnic background**  **White**  **Mixed**  **Asian**  **Black** | 11,133 (93%)  113 (1%)  393 (3%)  357 (3%) | 7,844 (89%)  121 (1%)  298 (3%)  601 (7%) | 18,977 (91%)  234 (1%)  691 (3%)  958 (5%) | | 52,829 (98%)  247 (1%)  389 (1%)  233 (0,4%) | | 10,527 (97%)  89 (1%)  109 (1%)  116 (1%) | | 63,356 (98%)  336 (0.5%)  498 (1%)  349 (0.5%) |
| **Alcohol consumption at recruitment**  **Never**  **Previous**  **Current**  **Unknown or missing** | 656 (5%)  426 (4%)  10,907 (91%)  7 (0.1%) | 630 (7%)  320 (4%)  7,905 (89%)  9 (0.1%) | 1,286 (6%)  746 (4%)  18,812 (90%)  16 (0.1%) | | 1,880 (4%)  1,392 (3%)  50,408 (93%)  18 (0.1%) | | 363 (3%)  374 (3%)  10,099 (93%)  5 (<0.1%) | | 2243 (3%)  1,766 (3%)  60,507 (94%)  23 (0.1%) |
| **Smoking status at recruitment**  **Never**  **Previous**  **Current**  **Unknown or missing** | 6,804 (57%)  3,615 (30%)  1,542 (13%)  35 (0.3%) | 5,019 (57%)  2,413 (27%)  1,407 (16%)  25 (0.3%) | 11,823 (57%)  6,028 (29%)  2,949 (14%)  60 (0.3%) | | 34,120 (64%)  16,736 (31%)  2,764 (5%)  78 (0.1%) | | 6,244 (58%)  3,763 (35%)  817 (7%)  17 (0.2%) | | 40,364 (62%)  20,499 (32%)  3,581 (6%)  95 (0.1%) |
| **Parity**  **0 child**  **1 child**  **2 children or more** | 2,424 (20%)  1,656 (14%)  7,916 (66%) | 1,882 (21%)  1,258 (14%)  5,724 (65%) | 4,306 (21%)  2,914 (14%)  13,640 (65%) | | 12,051(22%)  6,686 (13%)  34,961 (65%) | | 2,570 (24%)  1,402 (13%)  6,869 (63%) | | 14,621 (23%)  8,088 (12%)  41,830 (65%) |
| **Age at first live birth**  **< 35 or = 35**  **> 35**  **Nullipara**  **Unknown or missing** | 7,763 (65%)  136 (1%)  2,424 (20%)  1,673 (14%) | 5,576 (63%)  135 (1%)  1,882 (21%)  1,271 (14%) | 13,339 (64%)  271 (1%)  4,306 (21%)  2,944 (14%) | | 33,899 (63%)  1,051 (2%)  12,051(22%)  6,697 (13%) | | 6,629 (61%)  237 (2%)  2,570 (24%)  1,405 (13%) | | 40,528 (63%)  1,288 (2%)  14,621 (23%)  8,102 (12%) |
| **Age at menarche**  **<12 or = 12 years**  **>14 years**  **Unknown or missing** | 4,485 (37%)  7,164 (60%)  347 (3%) | 3,186 (36%)  5,433 (61%)  245 (3%) | 7,671 (38%)  12,597 (60%)  592 (3%) | | 20,755 (39%)  31,665 (59%)  1,278 (2%) | | 4,216 (39%)  6,437 (59%)  188 (2%) | | 24,971 (39%)  38,102 (59%)  1,466 (2%) |
| **Menopause at recruitment**  **Yes**  **No**  **Unknown or missing** | 4,267 (36%)  5,645 (47%)  2,084 (17%) | 3,565 (40%)  3,756 (42%)  1,543 (17%) | 9,401 (38%)  7,832 (45%)  3,627 (17%) | | 13,717 (26%)  31,904 (59%)  8,077 (15%) | | 2,762 (26%)  6,430 (59%)  1,649 (15%) | | 16,479 (26%)  38,334 (59%)  9,726 (15%) |
| **Age at menopause (n (%))**  **< 51 or = 51 years**  **> 51**  **Premenopausal or unknown**  **Unknown or missing** | 3,549 (30%)  1,867 (16%)  5,645 (47%)  935 (8%) | 2,508 (28%)  1,106 (12%)  3,756 (42%)  1,494 (17%) | 6,057 (29%)  2,973 (14%)  7,832 (45%)  3,998 (12%) | | 18,003 (33%)  12,327 (23%)  13,717 (26%)  9,651 (18%) | | 3,643 (34%)  2,496 (23%)  2,762 (25%)  1,940 (18%) | | 21,646 (36%)  14,823 (26%)  16,479 (26%)  11,591 (18%) |
| **Use of contraception pill at recruitment**  **Ever**  **Never**  **Unknown or missing** | 1,675 (14%)  10,293 (86%)  28 (0.2%) | 1,249 (14%)  7,592 (86%)  23 (0.3%) | 2,904 (14%)  17,885 (86%)  51 (0.2%) | | 7,852 (15%)  45,781 (85%)  65 (0.1%) | | 1,356 (13%)  9,473 (87%)  12 (0.1%) | | 9,208 (14%)  55,254 (86%)  77 (0.1%) |
| **Use of HRT at recruitment**  **Ever**  **Never**  **Unknown or missing** | 8,391 (70%)  3,550 (29%)  55 (1%) | 6,434 (73%)  2,392 (27%)  38 (0.4%) | 14,825 (71%)  5,942 (29%)  93 (0.4%) | | 34,608 (64%)  19,001 (35%)  89 (0.2%) | | 7,055 (65%)  3,769 (35%)  17 (0.2%) | | 41,663 (65%)  22,770 (35%)  106 (0.2%) |
| **Family history of breast cancer (mother, siblings) at recruitment**  **Breast cancer in family history**  **Other cancer than breast cancer in family history**  **Missing or no cancer reported in family history** | 1254 (10%)  10,557 (88%)  185 (2%) | 856 (10%)  7,846 (88%)  162 (2%) | 2,110 (11%)  18,403 (88%)  347 (0.2%) | | 6,067 (11%)  46,948 (87%)  683 (1%) | | 1,272 (12%)  9,441 (87%)  128 (1%) | | 7,339 (11%)  56,389 (87%)  811 (1%) |
| **History of benign breast lesion at recruitment**  **Yes**  **Not reported** | 874 (7%)  11,122 (93%) | 589 (7%)  8,275 (93%) | 1,463 (7%)  19,397 (93%) | | 5,255 (10%)  48,443 (90%) | | 1,185 (11%)  9,656 (89%) | | 58,099 (90%)  6,440 (10%) |
| **Participation in mammography screening at recruitment)**  **Yes, in the last 6 years**  **Yes, but not since >6 years**  **No or not regularly** | 7,567 (63%)  338 (3%)  4,091 (34%) | 5,088 (57%)  258 (3%)  3,518 (40%) | 12,655 (61%)  596 (3%)  7,609 (36%) | | 41,065 (76%)  981 (2%)  11,652 (22%) | | 7,487 (69%)  294 (3%)  3,060 (28%) | | 1275 (2%)  48,552 (75%)  14,712 (23%) |
